# Fibroblast Growth Factor 23 is a strong independent marker of worse cardiovascular outcomes after an acute coronary syndrome

**DOI:** 10.1101/2023.05.03.23289489

**Authors:** Andrea Kallmeyer, Ana Pello, Ester Cánovas, Álvaro Aceña, María Luisa González-Casaus, Nieves Tarín, Carmen Cristóbal, Carlos Gutiérrez-Landaluce, Ana Huelmos, Aida Rodríguez-Valer, Óscar González-Lorenzo, Joaquín Alonso, Lorenzo López-Bescós, Jesús Egido, Ignacio Mahillo, Óscar Lorenzo, José Tuñón

## Abstract

**Background and aims:** This study aimed to assess the role of plasmatic fibroblast growth factor 23 (FGF23) as a prognostic marker after an acute coronary syndrome (ACS).

**Methods:** This prospective and multicentric study included 1,190 patients with ACS. FGF23 plasma levels and other components of mineral metabolism (calcidiol, parathormone [PTH], klotho, and phosphate), lipids, troponin, high-sensitivity C-reactive protein, N-terminal-pro-brain natriuretic peptide and estimated glomerular filtration rate (eGFR) were measured at discharge. The primary outcome was a combination of acute ischemic events, heart failure (HF) and death. Secondary outcomes were the separate components of the primary outcome.

**Results:** Median follow-up was 5.44 (3.03-7.46) years. 294 patients developed the primary outcome. Patients with FGF23 levels below the median were predominantly males, younger, and with lower load of cardiovascular risk factors. Calcidiol and PTH levels were lower among them. Multivariable analysis showed that FGF23 (HR 1.18 [1.08-1.29], p<0.001), calcidiol (HR 0.86 [0.74-1.00], p=0.046), previous CAD or cerebrovascular accidents, and hypertension were independent predictors of the primary outcome. The predictive power of FGF23 was homogeneous across different subgroups of population. FGF23 resulted an independent predictor of HF (HR 1.38 [1.22-1.57], p<0.001), and death (HR 1.21 [1.07-1.37], p=0.002), but not of acute ischemic events. According to renal function, FGF-23 was an independent predictor for the primary outcome in patients with estimated glomerular filtration rate (eGFR) above 60 ml/min/1.73m^2^.

**Conclusions:** FGF23 is a strong, independent predictor of HF and death among patients with ACS. This effect is homogeneous across different subgroups of population and not limited to patients with chronic kidney disease (CKD).

## INTRODUCTION

Mineral metabolism involves intricate and complex pathways which also participate in the pathogenesis of cardiovascular diseases. Fibroblast growth factor 23 (FGF23) has a key role in that metabolism and increasing evidence is emerging about its function as a marker of cardiovascular events in a myriad of cardiac settings, both in chronic kidney disease (CKD) patients, in whom it was first studied, and in patients with normal renal function. FGF-23, a phosphaturic hormone produced by bone cells, and its co-receptor klotho are the main players in the regulation of phosphate homeostasis, preventing its harmful retention, especially when kidney function declines [1]. Although it helps to maintain mineral homeostasis, excessive FGF23 increases have deleterious side effects. High FGF23 plasma levels have been linked to left ventricular hypertrophy, cardiac fibrosis, endothelial dysfunction, and vascular calcification [2–5], and, thus, they have been associated with an increase in mortality and cardiovascular events [6]. However, whereas high FGF23 levels can be considered a marker of cardiovascular risk among patients with CKD, we have also previously described how patients with normal estimated glomerular filtration rate (eGFR) may present with increased FGF23 plasmatic levels [7]. Moreover, we and others have demonstrated that increased levels of FGF-23 may predict the development of cardiovascular events in patients with stable coronary artery disease (CAD) [6,8]. Furthermore, paracrine mechanisms involving FGF23 and myofibroblasts crosstalk in the context of cardiac ischemia have been described [9] and, moreover, the prognostic role of FGF23 has been studied in other cardiac conditions, such as, heart failure (HF) with preserved ejection fraction and atrial fibrillation [10].

Additionally, recent data from a clinical trial have shown that FGF23 plasmatic levels act as an independent predictor of adverse outcomes in patients who have had a recent Acute Coronary Syndrome (ACS) [11]. However, in that paper, plasmatic levels of the remaining components of mineral metabolism (calcidiol, parathormone [PTH], phosphate, and the soluble form of the FGF23 receptor named Klotho) were not explored. These findings are relevant, as we had previously demonstrated that there are significant interactions between the prognostic power of some of those components [8].

In this study we analyzed the prognostic role of FGF23 and the other from the Biomarkers in Acute Coronary Syndrome AND Biomarkers in Acute Myocardial Infarction (BACS&BAMI) studies.

## METHODS

### Patients

The BACS & BAMI studies included patients admitted to five hospitals in Madrid with either non-ST elevation acute coronary syndrome (NSTEACS) or ST-elevation myocardial infarction (STEMI). Inclusion and exclusion criteria have been defined previously [12].

Between July 2006 and June 2014, 2,740 patients were discharged from the study hospitals with a diagnosis of NSTEACS or STEMI. 1,483 patients were excluded due to the following prespecified criteria: age over 85 years (16.4%), presence of disorders or toxic habits limiting survival (29.8%), impossibility to perform cardiac revascularization (9.6%), coexistence of other significant cardiopathy (5.7%), impossibility to perform follow-up (16.3%), clinical instability beyond the sixth day after the index event (10.9%), refusal to participate in the study (1.5%), and impossibility of the investigators to include them (9.8%). From the included 1,257 patients, 1,230 completed the follow-up. From these, 1,190 had an adequate assessment of the components of mineral metabolism. On admission, clinical variables were recorded, and plasma was taken for analysis. Last follow-up visits were carried out in June 2016.

### Ethics Statement

The research protocol conformed to the ethical guidelines of the 1975 *Declaration of Helsinki* as reflected in a priori approval by the human research committees of the institutions participating in this study: Fundación Jiménez Díaz, Hospital Fundación Alcorcón, Hospital de Fuenlabrada, Hospital Universitario Puerta de Hierro Majadahonda, and Hospital Universitario de Móstoles. All patients signed informed consent documents. All patients signed informed consent documents. Date of approval by the Ethics Committee was 24th April 2007 (act number 05-07).

### Study Design

At baseline, clinical variables were recorded, and twelve-hour fasting venous blood samples were withdrawn and collected in EDTA. Blood samples were centrifuged at 2500 g for 10 minutes and plasma was stored at –80°C. Patients were seen every year at their hospitals. At the end of follow-up, the medical records were reviewed, and patient status was confirmed by telephone contact.

The primary outcome was the combination of acute ischemic events (STEMI, non-STEMI, unstable angina, Transient Ischemic Attack, and stroke), HF (both new onset HF and decompensated HF), and all-cause mortality. Secondary outcomes were the components of the primary outcome: acute ischemic events, HF, and death. NSTEACS was defined as rest angina lasting more than 20 minutes in the previous 24 hours, or new-onset class III-IV angina, along with transient ST depression or T wave inversion in the electrocardiogram considered diagnostic by the attending cardiologist and/or troponin elevation. STEMI was defined as symptoms compatible with angina lasting more than 20 minutes and ST elevation in at least two adjacent leads in the electrocardiogram without response to nitro-glycerine, and troponin elevation. A previous AMI was diagnosed in the presence of new pathological Q waves in the electrocardiogram along with a concordant new myocardial scar identified either by echocardiography or nuclear magnetic resonance imaging [13]. Stroke was defined as rapid onset of a neurologic defect attributable to a focal vascular territory lasting more than 24 hours or confirmed by new cerebral ischemic lesions on imaging studies. Transient Ischemic Attack was defined as a transient stroke with signs and symptoms resolved within the first 24 hours and without cerebral acute ischemic lesions at imaging techniques.

Although all events were recorded for each case, patients were excluded from the Cox regression analysis after the first event. Then, although the total number of events is also described, patients that had had more than one event were computed only once for these analyses. Patient’s follow-up was rigorous, made by clinical visits and also by telephone contact and hospital records reviews.

### Biomarker and Analytical Studies

Plasma determinations were performed at the laboratory of Mineral Metabolism, in Hospital La Paz, and the laboratories of Vascular Pathology and Biochemistry at Fundación Jiménez Díaz. The investigators who performed the laboratory studies were unaware of clinical data. Plasma calcidiol levels were quantified by chemiluminescent immunoassay (CLIA) on the LIAISON XL analyzer (LIAISON 25OH-Vitamin D total Assay DiaSorin, Saluggia, Italy), FGF-23 was measured by an enzyme-linked immunosorbent assay which recognizes epitopes within the carboxyl-terminal portion of FGF-23 (Human FGF23, C-Term, Immutopics Inc, San Clemente, CA), klotho levels by ELISA (Human soluble alpha klotho assay kit, Immuno-Biological Laboratories Co., Hokkaido, Japan), intact PTH was analyzed by a second-generation automated chemiluminescent method (Elecsys 2010 platform, Roche Diagnostics, Mannheim, Germany), and phosphate was determined by an enzymatic method (Integra 400 analyzer, Roche Diagnostics, Mannheim, Germany). N-Terminal pro-brain natriuretic peptide (NT-proBNP) levels were assessed by immunoassay (VITROS, Orthoclinical Diagnostics, Raritan, NJ, U.S.A.), high-sensivity C-reactive protein (hs-CRP) by latex-enhanced immunoturbidimetry (ADVIA 2400 Chemistry System, Siemens, Munich, Germany), and troponin by immunometric immunoassay with a mice biotin-monoclonal antibody and a luminescent reaction (Ortho Clinical Diagnostics Vitros XT 7600, Raritan, NJ, U.S.A.). Lipids, glucose and creatinine determinations were performed by standard methods (ADVIA 2400 Chemistry System, Siemens, Munich, Germany). The eGFR was calculated using the Chronic Kidney Disease Epidemiology Collaboration equation.

### Statistical Analysis

Quantitative data following a normal distribution are presented as mean ± standard deviation, and those with a not normal distribution are displayed as median (interquartile range). Qualitative variables are presented as percentages.

Differences in baseline data of patients according to baseline FGF23 values above the median or not, were assessed using Chi-squared or Fisher exact test for qualitative data. For quantitative variables, a t-Student test was performed for those following a normal distribution, and the Mann-Whitney test was used in those not normally distributed.

Univariable Cox regression was performed to analyse which variables were associated with the development of the outcomes. Then, multivariable regression analysis was carried out including those variables that achieved statistical significance at univariable analyses. Kaplan-Meier curves were traced showing the development of the different outcomes according to baseline FGF23 levels above the median or not. Interaction tests were performed to look for differences in the predictive power of FGF23 over median in different subgroups. Cox uni-and multivariable analyses were repeated dividing the population between those with eGFR<60 ml/min/1.73 m^2^ or not.

Analyses were performed with SPSS 20.0 (SPSS Inc., New York), and were considered significant when ‘‘p’’ was lower than 0.05 (two-tailed).

## RESULTS

### Characteristics of the population

We divided our study population according to FGF23 plasma levels above or under the median (110 RU/mL). Median time for blood extraction from admission was 4 (2-5) days. Median follow-up was 5.44 (3.03-7.46) years. Patients in the FGF23≤110 RU/mL were less frequently women, significantly younger, and had a lower body mass index. Also, they were less frequently diabetic, and had lower rates of hypertension.

Previous CAD was significantly less frequent in the FGF23≤110 RU/mL, but complete revascularization at the index event was significantly higher among them. More patients in the low FGF23 group were smokers. Atrial fibrillation rates were scarce, but higher among in the high FGF23 group. No differences were shown on left ventricular ejection fraction (LVEF)<40%. In the high FGF23 group eGFR resulted lower. PTH and FGF23 levels were significantly lower in the FGF23≤110 RU/mL group. About pharmacological treatments, anticoagulant therapy, insulin, diuretics, and aldosterone receptor blockers were used less often in the FGF23≤110 RU/mL, while betablockers were used more frequently (table 1).

**Table 1:**
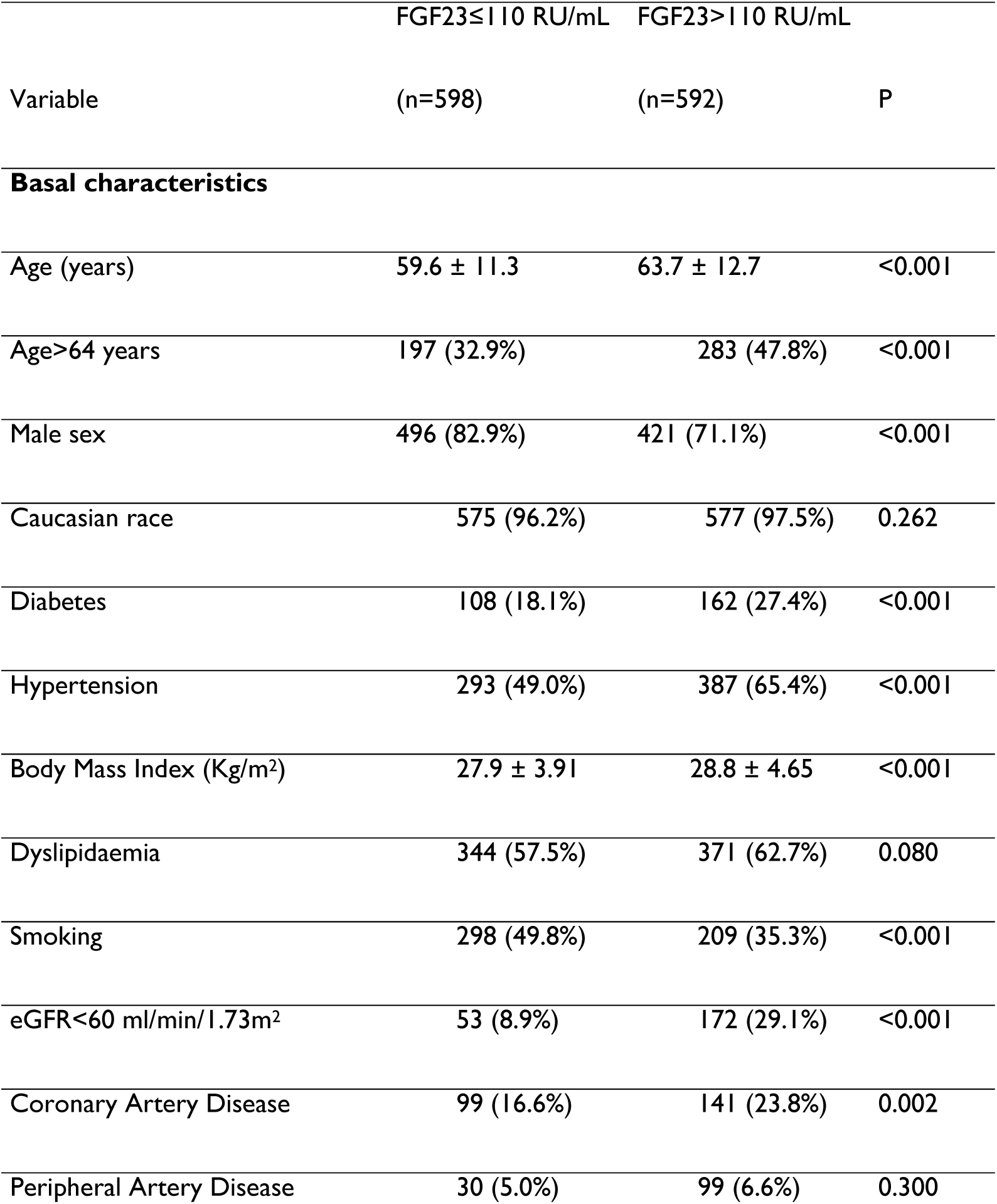
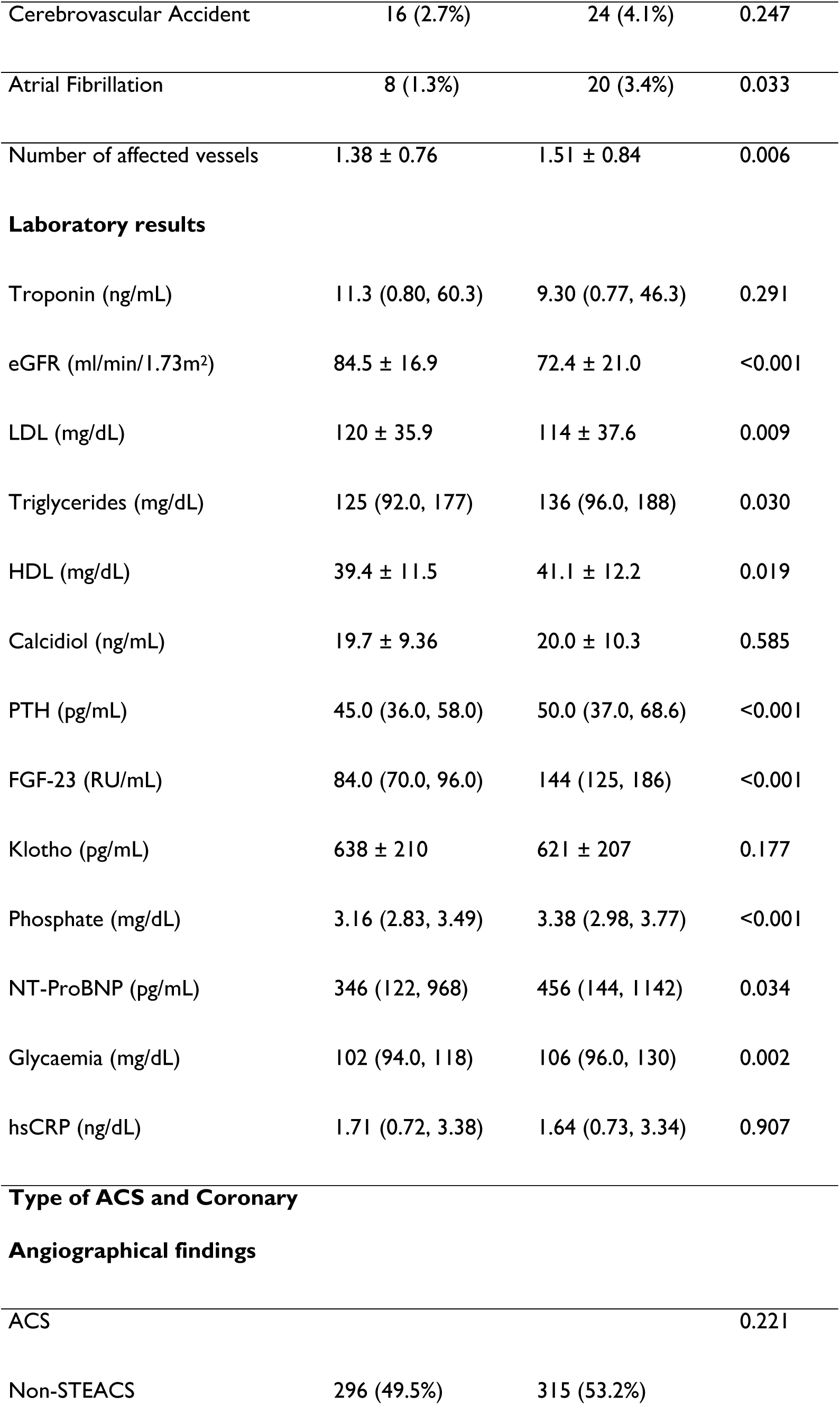
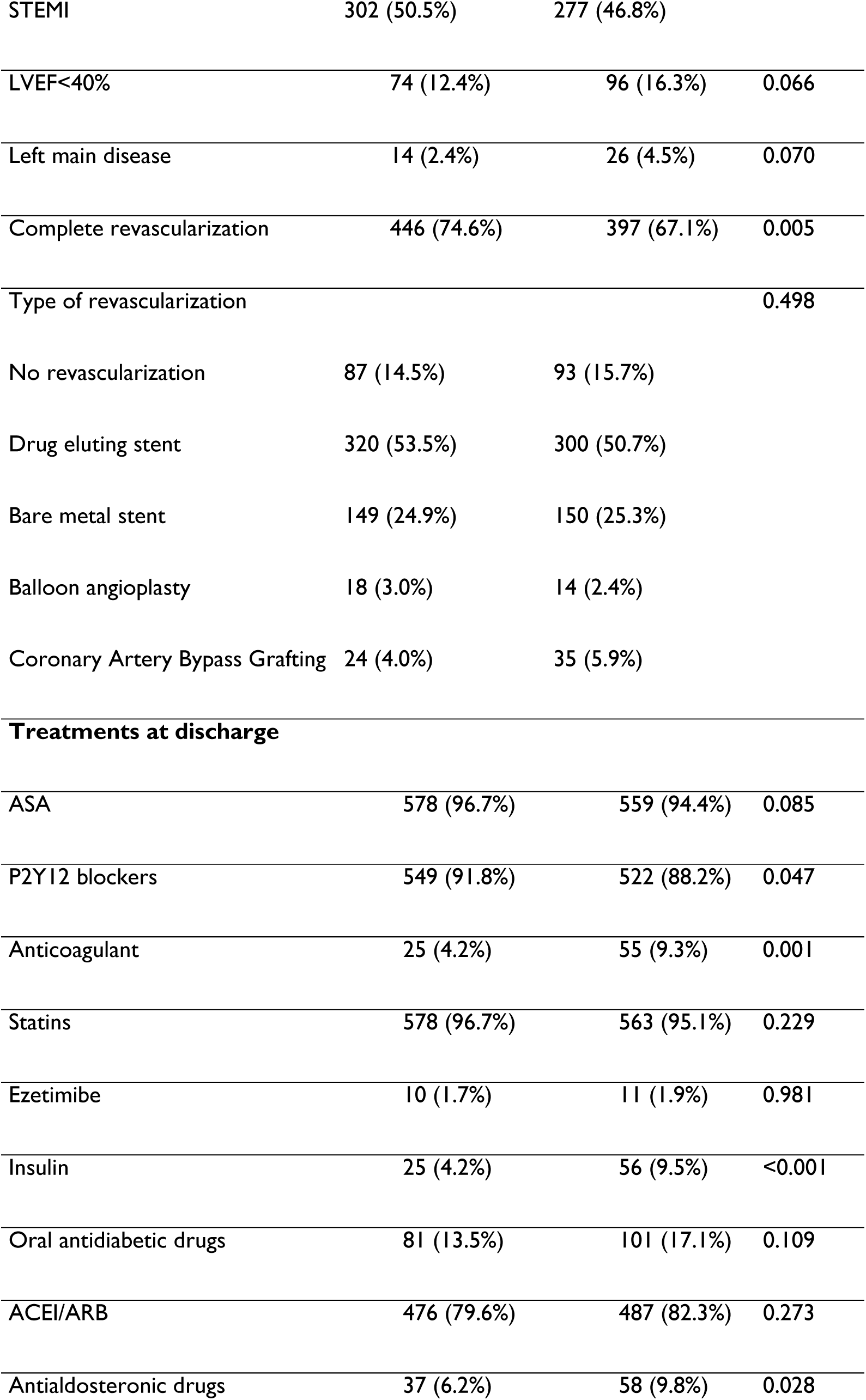
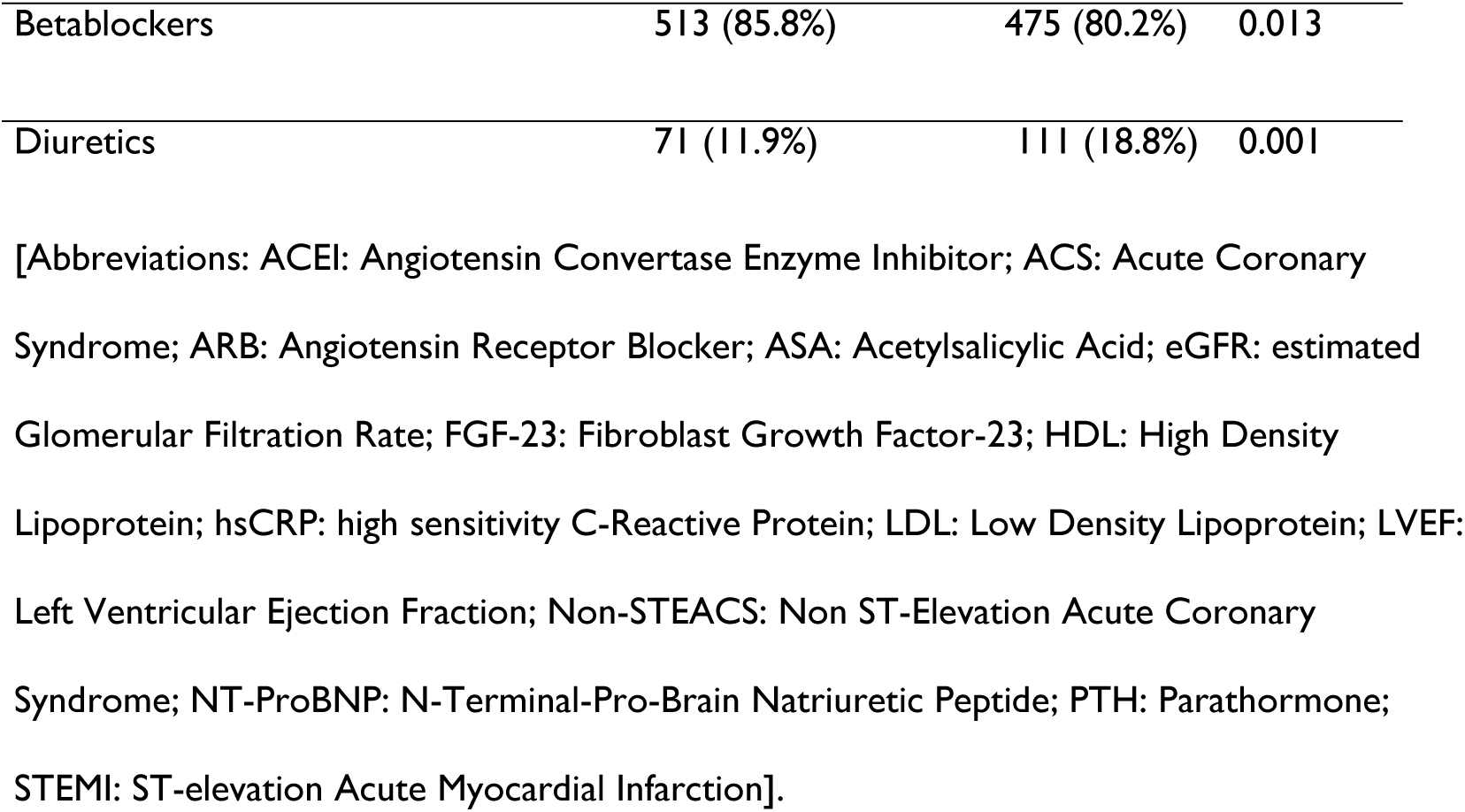
Baseline data dividing the population according to median FGF-23 values.

|  | FGF23≤110 RU/mL | FGF23>110 RU/mL |  |
| --- | --- | --- | --- |
| Variable | (n=598) | (n=592) | P |
| **Basal characteristics** |  |  |  |
| Age (years) | 59.6 ± 11.3 | 63.7 ± 12.7 | <0.001 |
| Age>64 years | 197 (32.9%) | 283 (47.8%) | <0.001 |
| Male sex | 496 (82.9%) | 421 (71.1%) | <0.001 |
| Caucasian race | 575 (96.2%) | 577 (97.5%) | 0.262 |
| Diabetes | 108 (18.1%) | 162 (27.4%) | <0.001 |
| Hypertension | 293 (49.0%) | 387 (65.4%) | <0.001 |
| Body Mass Index (Kg/m<sup>2</sup>) | 27.9 ± 3.91 | 28.8 ± 4.65 | <0.001 |
| Dyslipidaemia | 344 (57.5%) | 371 (62.7%) | 0.080 |
| Smoking | 298 (49.8%) | 209 (35.3%) | <0.001 |
| eGFR<60 ml/min/1.73m<sup>2</sup> | 53 (8.9%) | 172 (29.1%) | <0.001 |
| Coronary Artery Disease | 99 (16.6%) | 141 (23.8%) | 0.002 |
| Peripheral Artery Disease | 30 (5.0%) | 99 (6.6%) | 0.300 |
| Cerebrovascular Accident | 16 (2.7%) | 24 (4.1%) | 0.247 |
| Atrial Fibrillation | 8 (1.3%) | 20 (3.4%) | 0.033 |
| Number of affected vessels | 1.38 ± 0.76 | 1.51 ± 0.84 | 0.006 |
| Laboratory results |  |  |  |
| Troponin (ng/mL) | 11.3 (0.80, 60.3) | 9.30 (0.77, 46.3) | 0.291 |
| eGFR (ml/min/1.73m<sup>2</sup>) | 84.5 ± 16.9 | 72.4 ± 21.0 | <0.001 |
| LDL (mg/dL) | 120 ± 35.9 | 114 ± 37.6 | 0.009 |
| Triglycerides (mg/dL) | 125 (92.0, 177) | 136 (96.0, 188) | 0.030 |
| HDL (mg/dL) | 39.4 ± 11.5 | 41.1 ± 12.2 | 0.019 |
| Calcidiol (ng/mL) | 19.7 ± 9.36 | 20.0 ± 10.3 | 0.585 |
| PTH (pg/mL) | 45.0 (36.0, 58.0) | 50.0 (37.0, 68.6) | <0.001 |
| FGF-23 (RU/mL) | 84.0 (70.0, 96.0) | 144 (125, 186) | <0.001 |
| Klotho (pg/mL) | 638 ± 210 | 621 ± 207 | 0.177 |
| Phosphate (mg/dL) | 3.16 (2.83, 3.49) | 3.38 (2.98, 3.77) | <0.001 |
| NT-ProBNP (pg/mL) | 346 (122, 968) | 456 (144, 1142) | 0.034 |
| Glycaemia (mg/dL) | 102 (94.0, 118) | 106 (96.0, 130) | 0.002 |
| hsCRP (ng/dL) | 1.71 (0.72, 3.38) | 1.64 (0.73, 3.34) | 0.907 |
| Angiographical findings |  |  |  |
| ACS |  |  | 0.221 |
| Non-STEACS | 296 (49.5%) | 315 (53.2%) |  |
| STEMI | 302 (50.5%) | 277 (46.8%) |  |
| LVEF<40% | 74 (12.4%) | 96 (16.3%) | 0.066 |
| Left main disease | 14 (2.4%) | 26 (4.5%) | 0.070 |
| Complete revascularization | 446 (74.6%) | 397 (67.1%) | 0.005 |
| Type of revascularization |  |  | 0.498 |
| No revascularization | 87 (14.5%) | 93 (15.7%) |  |
| Drug eluting stent | 320 (53.5%) | 300 (50.7%) |  |
| Bare metal stent | 149 (24.9%) | 150 (25.3%) |  |
| Balloon angioplasty | 18 (3.0%) | 14 (2.4%) |  |
| Coronary Artery Bypass Grafting | 24 (4.0%) | 35 (5.9%) |  |
| **Treatments at discharge** |  |  |  |
| ASA | 578 (96.7%) | 559 (94.4%) | 0.085 |
| P2Y12 blockers | 549 (91.8%) | 522 (88.2%) | 0.047 |
| Anticoagulant | 25 (4.2%) | 55 (9.3%) | 0.001 |
| Statins | 578 (96.7%) | 563 (95.1%) | 0.229 |
| Ezetimibe | 10 (1.7%) | 11 (1.9%) | 0.981 |
| Insulin | 25 (4.2%) | 56 (9.5%) | <0.001 |
| Oral antidiabetic drugs | 81 (13.5%) | 101 (17.1%) | 0.109 |
| ACEI/ARB | 476 (79.6%) | 487 (82.3%) | 0.273 |
| Antialdosteronic drugs | 37 (6.2%) | 58 (9.8%) | 0.028 |
| Betablockers | 513 (85.8%) | 475 (80.2%) | 0.013 |
| Diuretics | 71 (11.9%) | 111 (18.8%) | 0.001 |

[Abbreviations: ACEI: Angiotensin Convertase Enzyme Inhibitor; ACS: Acute Coronary Syndrome; ARB: Angiotensin Receptor Blocker; ASA: Acetylsalicylic Acid; eGFR: estimated Glomerular Filtration Rate; FGF-23: Fibroblast Growth Factor-23; HDL: High Density Lipoprotein; hsCRP: high sensitivity C-Reactive Protein; LDL: Low Density Lipoprotein; LVEF: Left Ventricular Ejection Fraction; Non-STEACS: Non ST-Elevation Acute Coronary Syndrome; NT-ProBNP: N-Terminal-Pro-Brain Natriuretic Peptide; PTH: Parathormone; STEMI: ST-elevation Acute Myocardial Infarction].

### Independent predictors of the primary outcome

294 patients (24.7%) of the total population developed the primary outcome. Among them: 141 patients developed an ACS, 49 a cerebrovascular accident, 68 heart failure, and 115 died. Ten patients developed three different types of events, 59 had two different events, and the remaining patients developed a single event.

Univariable analysis showed that age, diabetes, hypertension, previous cardiovascular disorders, LVEF<40% and therapy with anticoagulant, insulin and antidiabetic drugs, mineralcorticoid receptor antagonist (MRA) or diuretics, among other factors, were positively associated with the development of the primary outcome. Other factors, such as eGFR, complete revascularization and the use of acetylsalicylic acid (ASA), P2Y12 blockers, statins, and betablockers, showed an inverse association with the outcome (table A from Supplemental Material).

Multivariable analysis (table 2) showed that high FGF23, CAD or stroke, age, hypertension, LVEF<40%, diuretics, and insulin were independently associated with the outcome. Calcidiol levels and the use of betablockers and modulators of renin-angiotensin system were inversely and independently related to the outcome. No interaction was found between FGF23 and calcidiol (p 0.71).

**Table 2:**
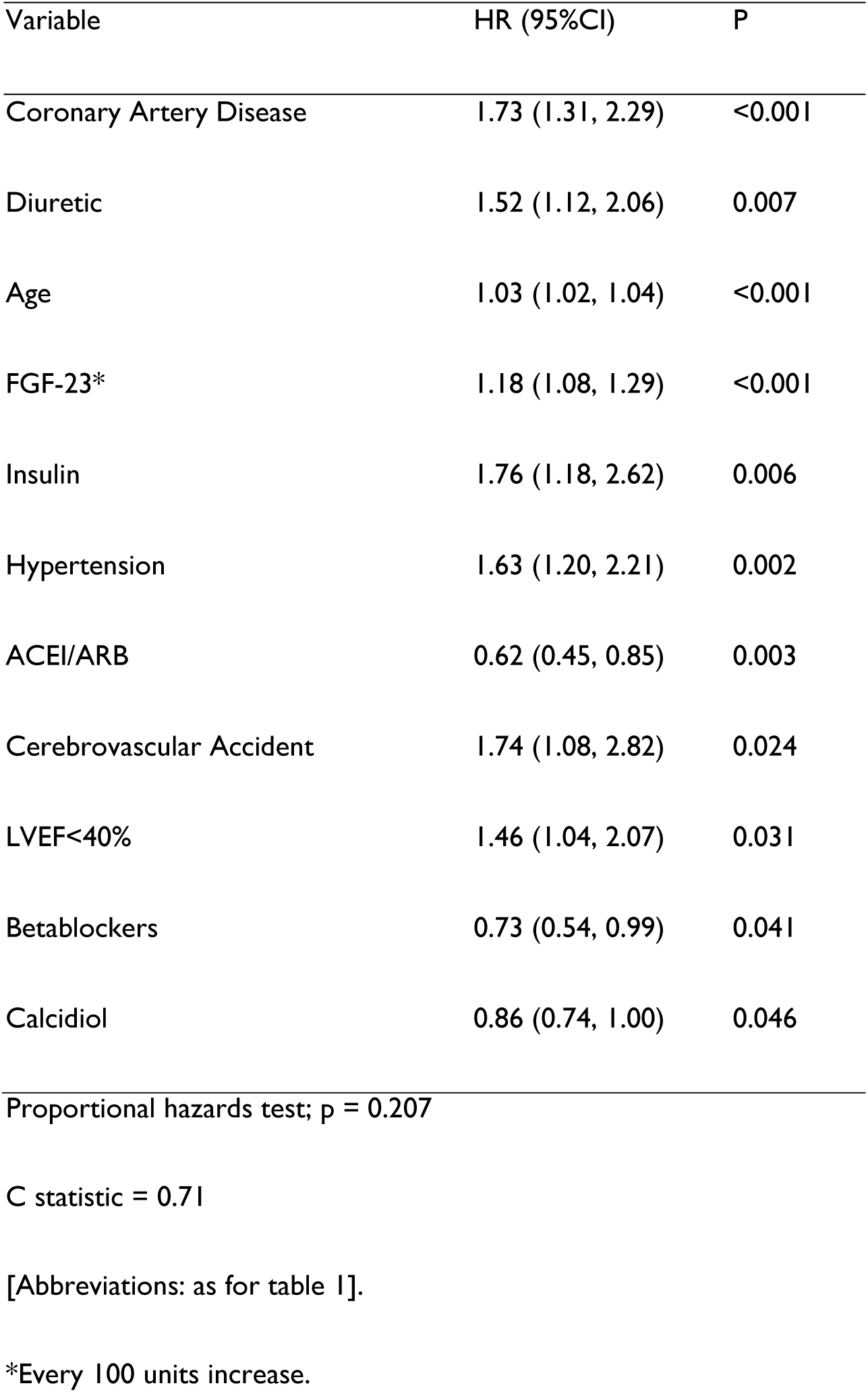
Multivariable Cox regression analysis for the primary outcome of acute ischemic event, heart failure or death.

| Variable | HR (95%CI) | P |
| --- | --- | --- |
| Coronary Artery Disease | 1.73 (1.31, 2.29) | <0.001 |
| Diuretic | 1.52 (1.12, 2.06) | 0.007 |
| Age | 1.03 (1.02, 1.04) | <0.001 |
| FGF-23* | 1.18 (1.08, 1.29) | <0.001 |
| Insulin | 1.76 (1.18, 2.62) | 0.006 |
| Hypertension | 1.63 (1.20, 2.21) | 0.002 |
| ACEI/ARB | 0.62 (0.45, 0.85) | 0.003 |
| Cerebrovascular Accident | 1.74 (1.08, 2.82) | 0.024 |
| LVEF<40% | 1.46 (1.04, 2.07) | 0.031 |
| Betablockers | 0.73 (0.54, 0.99) | 0.041 |
| Calcidiol | 0.86 (0.74, 1.00) | 0.046 |

Proportional hazards test; p = 0.207
C statistic = 0.71
[Abbreviations: as for table 1].
\*Every 100 units increase.

### Independent predictors of the incidence of acute ischemic events

184 patients (15.5%) developed an acute ischemic event. Six developed both an ACS and a cerebrovascular accident (CVA), and the remaining subjects developed a single event. The univariable analysis is presented in table B (Supplemental Material).

Multivariable analysis (table 3) showed that no biomarker had an independent predictive power for the outcome, while previous CAD and CVA, and hypertension had an independent positive association with it. On the other hand, STEMI as the index event, eGFR, and angiotensine-convertase enzyme inhibitors/angiotensin II receptor blockers treatment showed an inverse relationship.

**Table 3:** Multivariable Cox regression analysis for the secondary outcome of acute ischemic events.

| Variable | HR (95%CI) | P |
| --- | --- | --- |
| Coronary Artery Disease | 1.78 (1.25, 2.53) | 0.001 |
| eGFR | 0.89 (0.82, 0.97) | 0.006 |
| STEMI | 0.69 (0.48, 0.99) | 0.043 |
| Hypertension | 1.61 (1.10, 2.36) | 0.014 |
| ACEI/ARB | 0.63 (0.43, 0.92) | 0.016 |
| Cerebrovascular Accident | 1.88 (1.01, 3.51) | 0.047 |

Proportional hazards test; p = 0.148
C statistic = 0.67
[Abbreviations: as for table 1].

### Independent predictors of the development of heart failure

68 patients (5.7%) developed HF (Supplemental Material, table C). At multivariable analysis, high FGF23 resulted an independent predictor of HF, as well as PTH, LVEF<40%, previous CVA, age, and treatment with antidiabetic drugs or MRA. HDL was inversely related to the outcome (table 4). No interactions were found among FGF-23 and PTH (p=0.988).

**Table 4:** Multivariable Cox regression analysis for the secondary outcome of heart failure.

| Variable | HR (95%CI) | P |
| --- | --- | --- |
| FGF23 | 1.45 (1.28, 1.65) | <0.001 |
| PTH | 1.06 (1.01, 1.12) | 0.032 |
| LVEFE<40% | 3.84 (1.98, 7.45) | <0.001 |
| Previous CVA | 3.73 (1.84, 7.56) | <0.001 |
| Age | 1.08 (1.05, 1.11) | <0.001 |
| Antidiabetic drugs | 2.9 (1.61, 5.22) | <0.001 |
| HDL | 0.73 (0.57, 0.93) | 0.012 |
| Antialdosteronic drugs | 2.44 (1.15, 5.15) | 0.019 |

Proportional hazards test; p = 0.042
C statistic = 0.88
[Abbreviations: as for table 1].

### Independent predictors of death

115 patients (9.7%) died. The cause of death was cardiovascular in 48 cases (4.0%), cancer in 18 (1.5%), infection in 14 (1.2%), unknown in 14 (1.2%), renal failure in three (0.3%), pancreatitis in two (0.2%), gastro-intestinal bleeding in two (0.2%), and other in 14 (1.2%). The univariable analysis is shown in table D (Supplemental Material). After multivariable analysis (table 5), FGF-23 and calcidiol, age, insulin, LVEF<40%, ASA, MRA, and LDL resulted independent predictors of death, being the latter two protective. No interactions were found among FGF-23 and calcidiol.

**Table 5:**
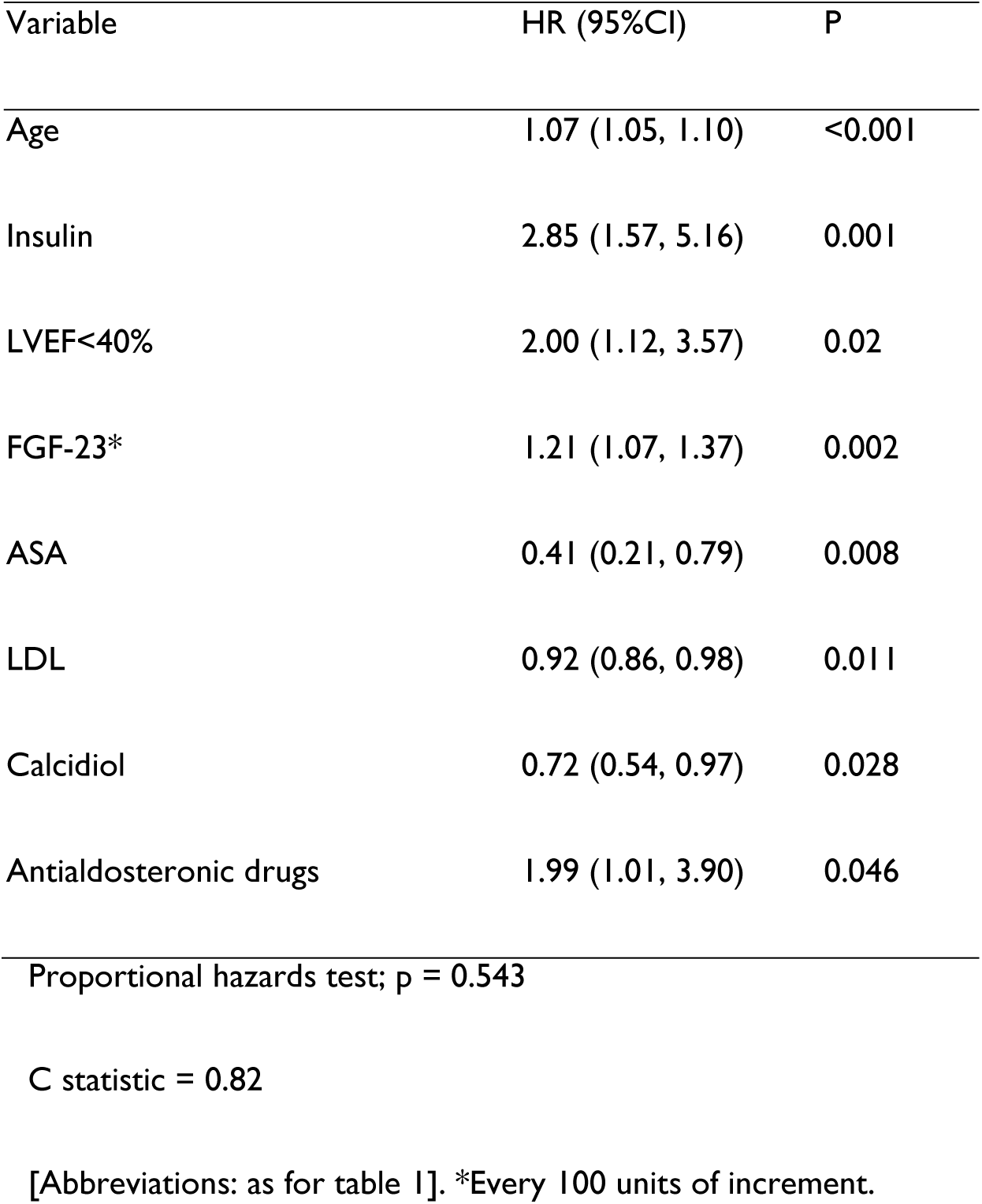
Multivariable Cox regression analysis for the secondary outcome of death.

### Prognostic power of FGF23 plasma levels above the median

Kaplan-Meier curves showed highly significant differences in patients with FGF23 levels above the median (110 RU/mL) about the primary outcome, HF, and death (Figure 1). We also performed Kaplan-Meier curves by dividing the population into two groups according to the FGF23 cut-off value of 92.8 RU/mL, which was found in the SOLID-TIMI trial to be the one that separated quartile 4 from quartiles 1-3 in that study [6]. The results were not more evident than using the median FGF23 cut-off. In fact, this threshold value represented the percentil 35^th^ in our population. In our study blood was withdrawn at discharge of the index event, whereas in the SOLID-TIMI trial it was taken about 15 days after the index event. Thus, we hypothesized that FGF23 levels could decrease after an ACS, which was confirmed in our series (Table 6).

**Figure 1:**
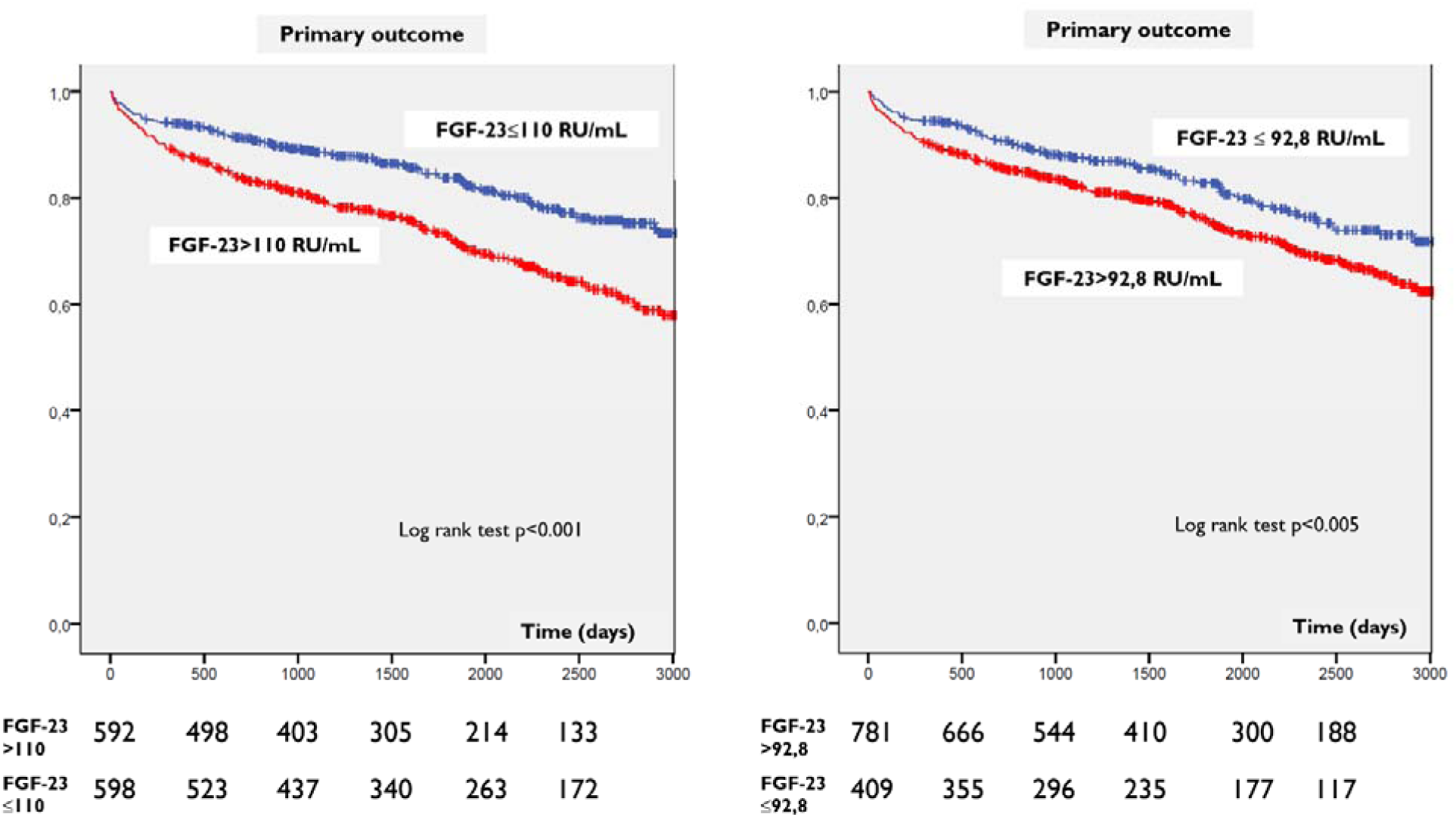
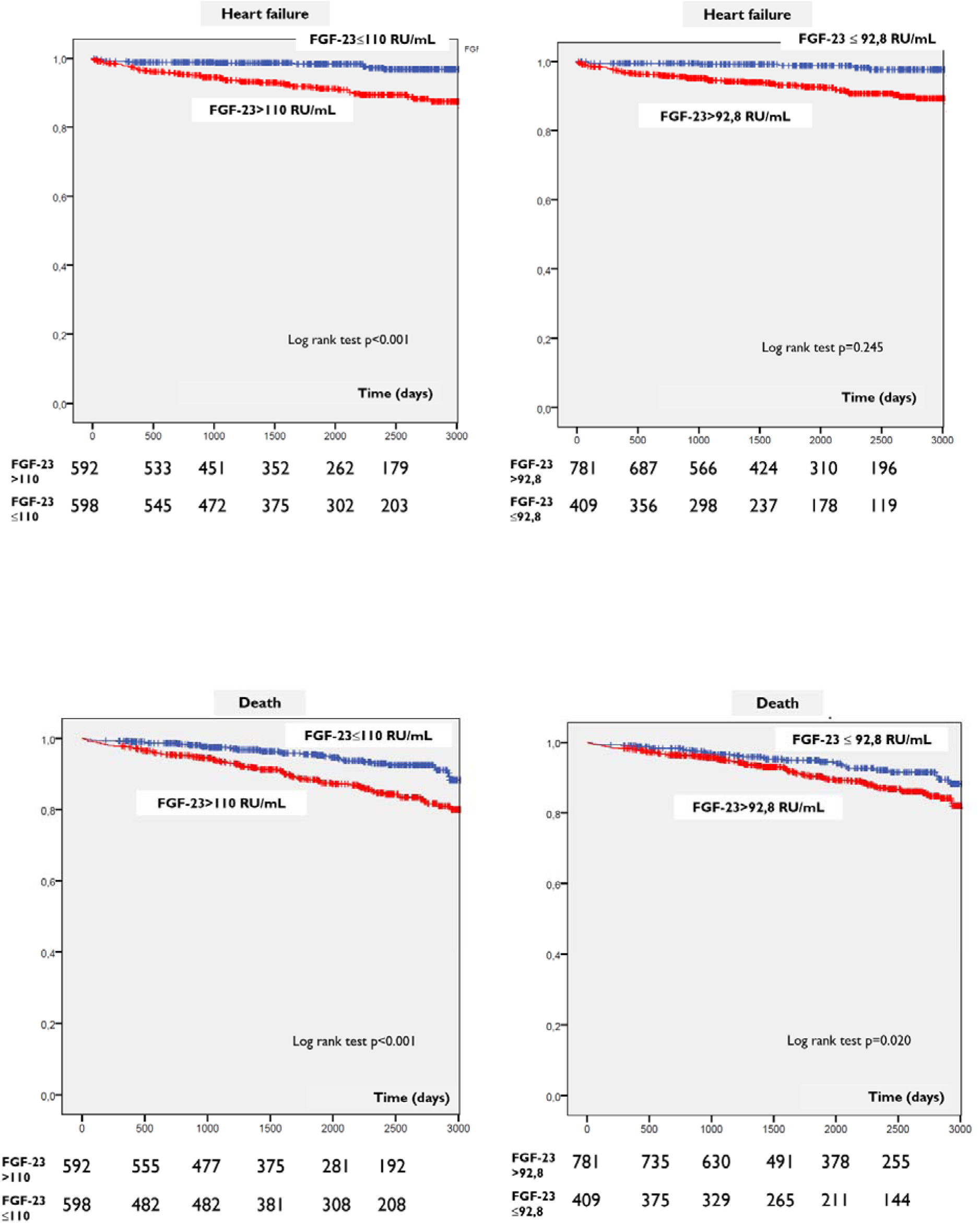
Kaplan-Meier curves showing the incidence of cardiovascular events once divided the population according to median FGF23 values with the cut-off values of our study (110 RU/mL) and the cut-off values of the SOLID-TIMI52 trial (92,8 RU/mL) [11].

**Table 6:**
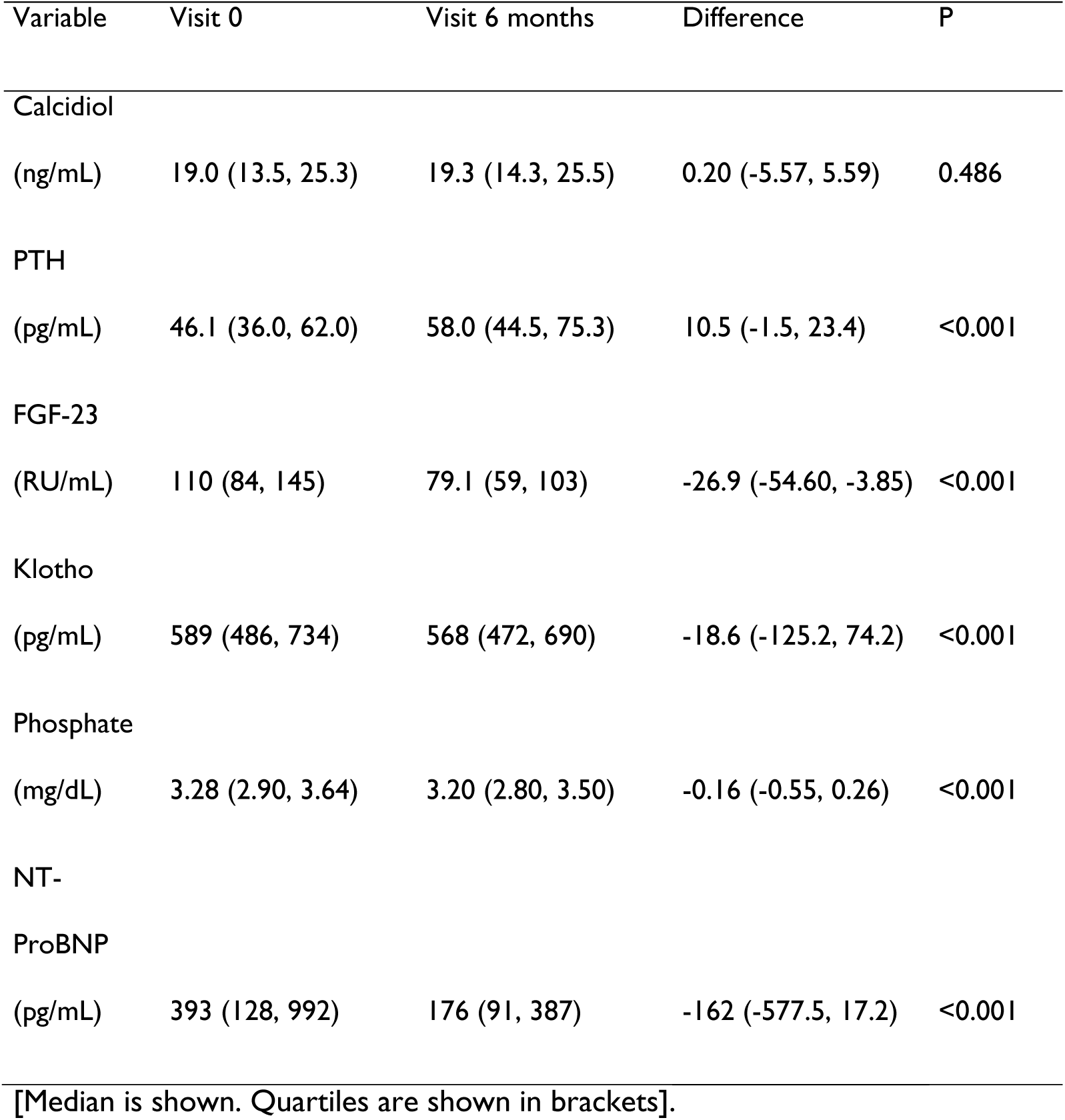
Comparison of plasma levels of mineral metabolism components at discharge and 6 months later.

| Variable | Visit 0 | Visit 6 months | Difference | P |
| --- | --- | --- | --- | --- |
| Calcidiol |  |  |  |  |
| (ng/mL) | 19.0 (13.5, 25.3) | 19.3 (14.3, 25.5) | 0.20 (-5.57, 5.59) | 0.486 |
| PTH |  |  |  |  |
| (pg/mL) | 46.1 (36.0, 62.0) | 58.0 (44.5, 75.3) | 10.5 (-1.5, 23.4) | <0.001 |
| FGF-23 |  |  |  |  |
| (RU/mL) | 110 (84, 145) | 79.1 (59, 103) | -26.9 (-54.60, -3.85) | <0.001 |
| Klotho |  |  |  |  |
| (pg/mL) | 589 (486, 734) | 568 (472, 690) | -18.6 (-125.2, 74.2) | <0.001 |
| Phosphate |  |  |  |  |
| (mg/dL) | 3.28 (2.90, 3.64) | 3.20 (2.80, 3.50) | -0.16 (-0.55, 0.26) | <0.001 |
| NT-ProBNP |  |  |  |  |
| (pg/mL) | 393 (128, 992) | 176 (91, 387) | -162 (-577.5, 17.2) | <0.001 |
| [Median is shown. Quartiles are shown in brackets]. |  |  |  |  |

### Homogeneity of the results obtained across different population subgroups for the studied outcomes

No significant differences were shown when the incidence of the primary outcome, HF, and death was assessed according to FGF23 levels across subgroups (figure 2).

**Figure 2:**
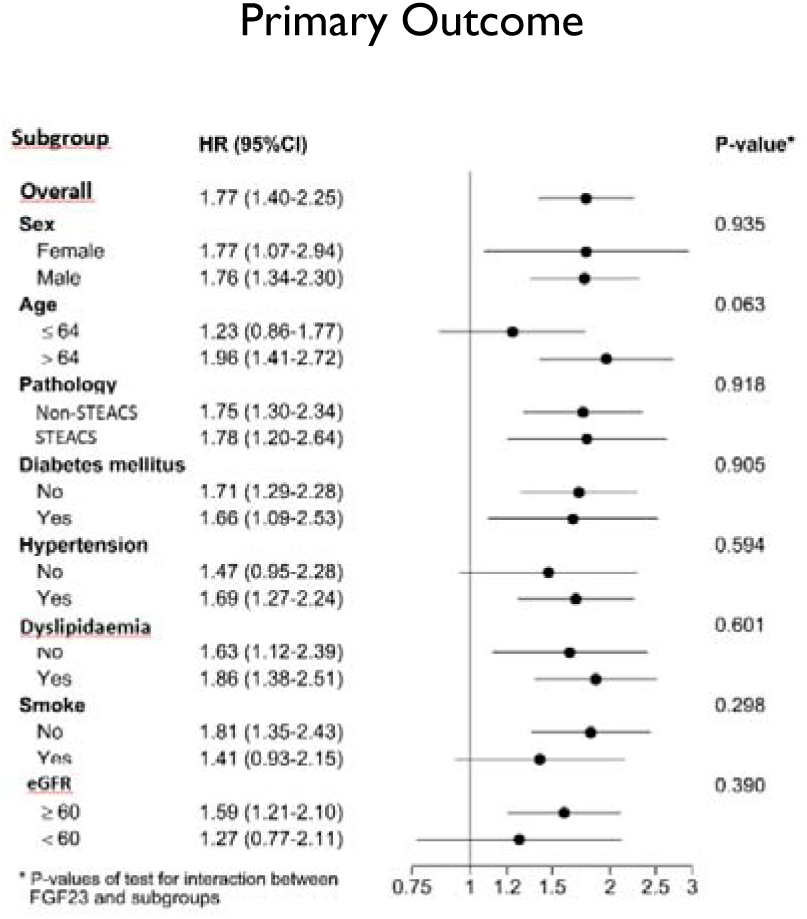
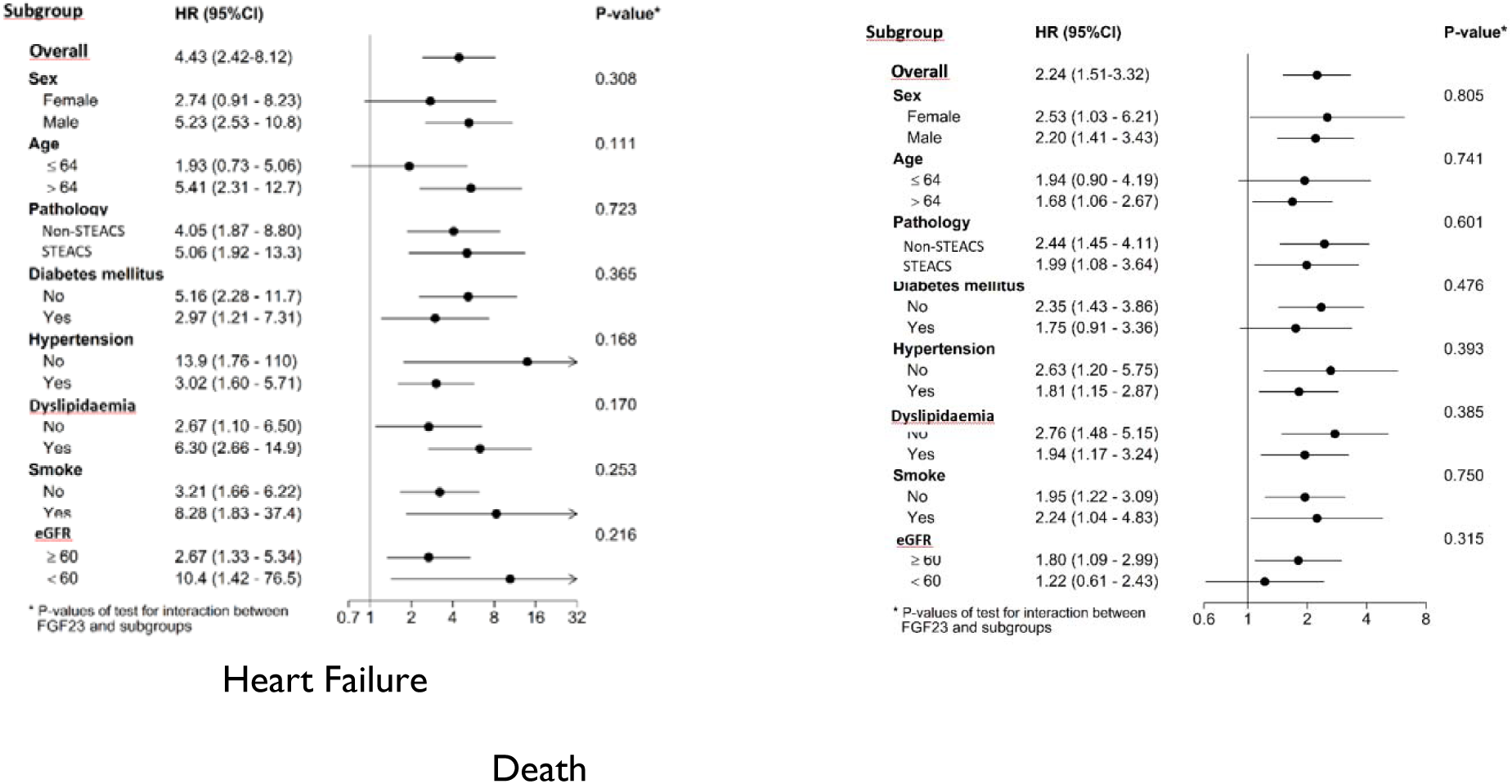
Forrest plots showing the prognostic value of FGF23 levels above the median for the primary and secondary outcomes across different population subgroups.

### Independent predictive power of FGF-23 for the primary outcome according to renal function

Two-hundred patients (18.9%) had an eGFR<60 ml/min/1.73 m^2^. In these subjects, no components of mineral metabolism were independently associated with the primary outcome. However, considering the group of eGFR ≥ 60 ml/min/1.73m^2,^ FGF23 was positively and independently associated with the primary outcome, while calcidiol was inversely related to it (table 7).

**Table 7:** Multivariable Cox regression analysis for the primary outcome according to eGFR

**eGFR ≥ 60 ml/min/1.73 m<sup>2</sup>:**
| Variable | HR (95%CI) | P |
| --- | --- | --- |
| Age | 1.02 (1.01-1.04) | 0.002 |
| FGF-23* | 1.16 (1.05-1.30) | 0.005 |
| Diuretics | 1.67 (1.12-2.48) | 0.012 |
| Hypertension | 1.49 (1.07-2.09) | 0.02 |
| Betablockers | 0.67 (0.47-0.96) | 0.031 |
| Insulin | 1.80 (1.01-3.21) | 0.044 |
| Calcidiol** | 0.82 (0.68-0.99) | 0.044 |

**eGFR < 60 ml/min/1.73 m<sup>2</sup>:**
| Variable | HR (95%CI) | P |
| --- | --- | --- |
| eGFR** | 0.74 (0.59-0.93) | 0.009 |
| Anticoagulant | 3.14 (1.69-5.84) | <0.001 |
| LDL** | 0.89 (0.84-0.95) | 0.004 |
| Age | 1.03 (1.00-1.06) | 0.030 |
| Preivous CVA | 2.16 (1.15-4.06) | 0.017 |

Proportional hazards test; p = 0.095
C statistic = 0.70
[Abbreviations: as for table 1] \* Every 100 units of increment. \*\* Every 10 units of increment.

## DISCUSSION

### Mineral metabolism

Mineral metabolism covers a complex network of metabolic pathways with reciprocal interactions between the functioning of bone, the kidneys, peripheral tissue, and the heart. This system maintains phosphate homeostasis in chronic kidney disease (CKD). From earliest stages of CKD, decrease in renal expression of klotho displays an adaptative increase of FGF23 as a response to maintain normophosphatemia. FGF23 exerts a phosphaturic effect and meanwhile prevents new phosphate entry from intestine and bone through the inhibitory action on vitamin D and PTH. However, in advanced stages of CKD, the hypovitaminosis D and its corresponding hypocalcemia lead on the contrary to the development of a secondary hyperparathyroidism [14]. Although FGF23 represents a compensatory mechanism, its excessive increase has harmful effects, as it has been shown that mineral metabolism disturbances play a significant role in the development of vascular diseases, being linked to higher mortality in CKD patients [15].

### Role of FGF-23 in cardiovascular disease

Abnormalities of mineral metabolism are not exclusive to CKD patients, as we have found previously that almost 20% of patients with stable CAD and eGFR≥90 ml/min/1.73 m^2^ have increased plasmatic concentrations of FGF23 and PTH [7]. In line with this, other situations different to CKD, such systemic inflammatory processes, diabetes, ACS, and shock, show high FGF23 levels [16]. In this regard, FGF23 seems to be involved in vascular and, specifically, myocardial damage [5,15], irrespective of the coexistence of traditional cardiovascular risks factors. Thus, it has been linked to many cardiac diseases, mainly HF, and acute [17,18,19] and chronic [6,7,19,20] CAD, through causing left ventricular hypertrophy (LVH) [5], cardiac fibrosis and ventricular remodelling. [16,21]. These effects have been called the paracrine expression of FGF23 and are responsible for the cardiac adaptation to its imbalance [22]. Fibroblast growth factor-23 is overexpressed in a mouse model of right ventricular pressure overload mainly when there is fibrosis accompanying ventricular hypertrophy, while its receptor FGFR1 is overexpressed in the fibroblasts [23]. In this way, FGF23 could act as a paracrine factor produced by cardiomyocytes, thus promoting fibrosis in adjacent fibroblasts. In addition, the pro-fibrotic effect of FGF23 on fibroblasts in culture shows a synergy with TGF-β1 [23], a well-known mediator of fibrosis. Animal models have also demonstrated that not only high FGF23 concentrations, but also the length of exposure to them are critical factors with regard to cardiac adaptations [17]. Furthermore, while the heart suffers as a result of those mineral metabolism changes, it is not only a passive spectator. On the contrary, severe cardiac conditions, such as cardiogenic shock, also increase the production of FGF23, creating a vicious circle of adverse adaptations [24]. No safe levels of FGF23 have been established, nor have any potential benefits of its blockade, as animal models have linked the latter to aortic calcification and higher mortality [24].

### Prognostic value of FGF23

In our study, patients with ACS showing FGF23 plasma levels above the median (110 RU/mL) developed the primary outcome more frequently during the follow-up than those with FGF23 levels below or equal to 110 RU/ml. High FGF23 levels proved to be an independent predictor of HF and death, but not of ischemic events, despite adjusting for a large set of variables, including cardiovascular risk factors, previous cardiovascular history, data from the index events, and medical therapy. Furthermore, adjusting for other components of mineral metabolism and more relevantly, for established biomarkers, such as NT-proBNP, did not find any limits on the prognostic value of FGF23. Moreover, NT-proBNP and hs-CRP plasma levels did not provide additional prognostic value and were not positive in the multivariable model. These results are consistent with previous studies in the field [25,26]. However, some differences should be noted. In a sample of 125 patients with acute HF, high FGF23 levels at discharge predicted a higher incidence of HF or death at one year of follow-up [25]. Also, in 1,099 patients with acute myocardial infarction, the prognostic value of 175 biomarkers was explored after a median follow-up of 6.6 years [26]. FGF23 was among those biomarkers predicting the outcome of total death, recurrent acute myocardial infarction, or HF hospitalization, although the number of clinical variables for which adjustment was performed was lower than in our study. A meta-analysis including more than 135,000 participants from the general population, showed that FGF23 is an independent predictor of the development of HF, stroke, and acute myocardial infarction [27]. In the community-based PREVEND (Prevention of Renal and Vascular End-Stage Disease) study, a higher FGF23 level was associated with the development of HF with reduced ejection fraction but not with the appearance of HF with preserved ejection fraction during follow-up [28].

Finally, in a secondary analysis from the Stabilization of Plaques Using Darapladib-Thrombolysis in Myocardial Infarction 52 (SOLID-TIMI 52) trial including 4,947 patients with ACS, FGF23 was found to be an independent predictor of cardiovascular death or hospitalization by HF [11]. However, some differences from our work should be noted. Firstly, in our study we analysed all components of mineral metabolism and not only FGF23 levels. Secondly, in the multivariable analysis we considered ongoing treatments at discharge, which may have an additional role in the prediction of recurrent events of interest. Finally, we performed FGF23 assessment at discharge after the index event, while in the SOLID-TIMI52, FGF23 determination was performed in the 30 days following the index event. This is more complicated than just assessing FGF23 during ACS admission. Interestingly, although they used the FGF23 concentration that separated quartile 4 from 1-3 as a cut-off, these values were clearly lower than the median from our study (92.8 mg/dl vs 110 mg/dl), and when we tried to use this cut-off in our population, the predictive value of FGF23 did not improve. We explained this difference as a result of the different timing of FGF23 measurement, and we confirmed in our own population that FGF23 levels decrease during the 6 months following the ACS. Therefore, it might not be the same to assess FGF23 levels during ACS than assessing it in the following weeks.

### Changes in FGF-23 plasma levels after an ACS

The variation of FGF-23 plasma levels in time raises some questions to be answered in the future. In a previous paper we analysed the prognostic value of components of mineral metabolism in 964 patients from this population, six months after the ACS, in other words, when the patients had stable CAD [29]. At this point, PTH but not FGF23 turned out to be an independent prognostic marker of acute ischemic events, HF, or death. Although the ultimate reason for this change is still unknown, we speculate that it may be related to the changes in the mineral metabolism that we observed the six-month period after the ACS. In this regard, in addition to the aforementioned decrease in FGF23 plasma levels, there is an increase of PTH levels and a decrease of soluble klotho and phosphate levels. These data, as well as the evidence of the interaction between the prognostic value of calcidiol and FGF23 that we had demonstrated previously [8], shows that a complete assessment of mineral metabolism may be necessary in order to answer the outstanding questions in this area in the future.

### Predictive power of FGF-23 across subgroups

Given the strong relationship between mineral metabolism and renal function, it was possible that the results observed were due to the prognostic power of FGF23 in patients with CKD. However, when we performed multivariable regression analysis by subgroups according to renal function, FGF23 was a strong, independent predictor of the primary outcome in patients with eGFR≥60 ml/min/1.73m^2^. In patients with lower eGFR it did not retain this predictive power, although this could be due to the low sample size of this subgroup. Similarly, when we performed subgroup analysis, no differences were evident for the predictive power of FGF23 according to sex, age, and the presence of hypertension or diabetes among other factors. These findings are similar to those reported in the sub-analysis of the SOLID-TIMI 52 trial [11].

### Limitations

As a main limitation, given the design of the study that required the collection of plasma for analysis at discharge no later than six days after admission, to achieve homogeneous results, we excluded 10.9% of ACS patients who did not meet this condition. This explains the low number of patients with LVEF<40% that were included. Therefore, these results should not be extrapolated to populations with a high percentage of patients with moderate or severe LV systolic dysfunction.

## CONCLUSIONS

Among a population of patients with ACS with average eGFR, FGF23 is a strong independent predictor of HF and death, even after adjustment of a large set of variables, as well as for the values of other components of mineral metabolism and NT-proBNP. This effect was homogeneous across different population subgroups and not limited to patients with CKD.

## Data Availability

Data are available

## ACKNOWLEDGEMENTS

We want to thank AdHoc Translations for their collaboration in the process of English language review of the manuscript.

## REFERENCES

1. Inoue Y, Segawa H, Kaneko I, Yamanaka S, Kusano K, Kawakami E, Furutani J, Ito M, Kuwahata M, Saito H, Fukushima N, Kato S, Kanayama H-O, Miyamoto K. Role of the vitamin D receptor in FGF23 action on phosphate metabolism. Biochem J 2005; 390: 325–331

2. Gutiérrez OM, Mannstadt M, Isakova T. Fibroblast growth factor 23 and mortality among patients undergoing haemodialysis. N Engl J Med 2008; 359(6): 584–592.

3. Lanlan L, Juan G. Intact Fibrobast Growth Factor 23 Regulates Chronic Kidney Disease-Induced Myocardial Fibrosis by Activating the Sonic Hedgehog Signaling Pathway. J Am Heart Assoc. 2022; 11: E026365. DOI: 10.1161/jaha.1222.026365.

4. Liu M, Li XC, Lu L, Cao Y, Sun RR, Chen S, Zhang PY. Cardiovascular disease and its relationship with chronic kidney disease. Eur Rev Med Pharmacol Sci. 2014; 18: 2918–2926.

5. Faul C, Amaral AP, Oskouei B, Hu M-C, Sloan A, Isakova T, Gutiérrez OM, Aguillon-Prada R, Lincoln J, Hare JM, Mundel P, Morales A, Scialla J, Fischer IM, Soliman EZ, Chen J, Go AS, Rosas SE, Nessel L, Townsend RR, Feldman HI, St John Sutton M, Ojo A, Gadegbeku C, di Marco GS, Reuter S, Kentrup D, Tiemann K, Brand M, Hill JA, Moe OW, Kuro-o M, Kusek JW, Keane MG, Wolf M. FGF23 induces left ventricular hypertrophy. J Clin Invest 2011; 121:4393–4408.

6. Parker BD, Schurgers LJ, Brandenburg VM, Christenson RH, Vermeer C, Ketteler M, Shlipak MG, Whooley MA, Ix JH. The associations of fibroblast growth factor 23 and uncarboxylated matrix Gla protein with mortality in coronary artery disease: the Heart and Soul Study. Ann Intern Med 2010; 152: 640–648.

7. González-Parra E, Aceña Á, Lorenzo Ó, Tarín N, González-Casaus ML, Cristóbal C, Huelmos A, Mahíllo-Fernández I, Pello AM, Carda R, Hernández-González I, Alonso J, Rodríguez-Artalejo F, López-Bescós L, Ortiz A, Egido J, Tuñón J. Important abnormalities of bone mineral metabolism are present in patients with coronary artery disease with a mild decrease of the estimated glomerular filtration rate. J Bone Miner Metab. 2016 Sep;34(5):587–98.

8. Tuñón J, Cristóbal C, Tarín N, Aceña Á, González-Casaus ML, Huelmos A, Alonso J, Lorenzo Ó, González-Parra E, Mahíllo-Fernández I, Pello AM, Carda R, Farré J, Rodríguez-Artalejo F, López-Bescós L, Egido J. Coexistence of low vitamin D and high fibroblast growth factor-23 plasma levels predicts an adverse outcome in patients with coronary artery disease.

9. Kazuhiro K, Yocichiro K, Uesegi K, Semba K, Urashima T, Akaike T, Minamisaawa S. Fibrosis growth factor 23 is a promoting factor for cardiac fibrosis in the presence of transforming growth factor-ß1. PLoS ONE. 2020; 15(4): e0231905. Doi: 10.1371/jurnal.pone.0231905.

10. Roy C, Lejeune S, Slimani A, de Meester C, Ahn SA, Rousseay MF, Mihaela A, Ginion A, Ferracin B, Pasquet A, Vancraeynest D, Beauloye C, Vanoverschelde JL, Horman S, Gruson D, Gerber BL, Pouleur AC. Fibroblast growth factor23: a biomarker of fibrosis and prognosis in heart failure with preserved ejection fraction. Esc Heart Failure. 2020; 7: 2494–2507. Doi: 10.1002/ehfj2.12816.

11. Bergmark BA, Udell JA, Morrow DA, Cannon CP, Steen DL, Jarolim P, Budaj A, Hamm C, Guo J, Im K, Kuder JF, Braunwald E, Sabatine MS, O’Donoghue ML. Association of Fibroblast Growth Factor 23 With Recurrent Cardiovascular Events in Patients After an Acute Coronary Syndrome: A Secondary Analysis of a Randomized Clinical Trial. JAMA Cardiol. 2018 Jun 1;3(6):473–480.

12. Tuñón J, Blanco-Colio L, Cristóbal C, Tarín N, Higueras J, Huelmos A, Alonso J, Egido J, Asensio D, Lorenzo Ó, Mahíllo-Fernández I, Rodríguez-Artalejo F, Farré J, Martín-Ventura JL, López-Bescós L. Usefulness of a combination of monocyte chemoattractant protein-1, galectin-3, and N-terminal probrain natriuretic peptide to predict cardiovascular events in patients with coronary artery disease. Am J Cardiol 2014;113:434–440.

13. Thygesen K, Alpert JS, Jaffe AS, et al. Fourth Universal Definition of Myocardial Infarction. ESC Scientific Document Group. Eur Heart J (2019) 40: 237.269.

14. Wolf M. Forging forward with 10 burning questions on FGF 23 in kidney disease. J Am Soc Nephrol. 2010; 21(9): 1427–35. doi: 10.1681/ASN.2009121293.

15. Vogt I, Haffner D, Leifheit-Nestler M. FGF 23 and Phosphate-Cardiovascular Toxins in CKD. Toxins 2019; 11: 167. doi: 10.3390/toxins11110647.

16. Rodelo-Haad C, Santamaria R, Muñoz-Castañeda JR, Pendón Ruiz de Mier MV, Martín-Malo A, Rodríguez M. FGF23, Biomarker or Target? Toxins 2019; 11: 175. doi: 10.3390/toxins11030175.

17. Pöss J, Mahfoud F, Seiler S. FGF23 is associated with increased disease severity and early mortality in cardiogenic shock. European Heart Journal Acute Cardiovascular Care 2013; 2(3): 211–218.

18. Reindl M, Reinstadler SJ, Freistritzer HJ. Fibroblast growth factor 23 as novel biomarker for early risk stratification after ST-elevation myocardial infarction. Heart 2017; 103 (11): 856–862.

19. Kurpas An, Supel K, Idzikkowska K, Zielinska M. FGF23: A Review of Its Role in Mineral Metabolism and Renal and Cardiovascular Disease. Hindawi Disease Markers 2021; doi.org/10.1155/2021/8821292.

20. Lutsey PL, Alonso A, Selvin E. Fibroblast growth factor 23 and incident coronary heart disease, heart failure and cardiovascular mortality: the atherosclerosis risk in communities’ study. Journal of American Heart Association 2014; 3(3).

21. Kawai M. The FGF23/Klotho axis in the regulation of mineral and metabolic homeostasis. Horm Mol Biol Clin Invest 2016; 28: 57–67. doi: 10.1515/hmbci-2015-0068.

22. Prié D. FGF23 and Cardiovascular Risk. Klotz communication 2020: Heart and Hormones. Annales d’ Endocrinologie 2020. doi.org/10.1016/j.ando.2020.03.007. FGF and CV risk.

23. Kuga K, Kusakari Y, Uesugi K, Semba K, Urashima T, Akaike T, Minamisawa S. Fibrosis growth factor 23 is a promoting factor for cardiac fibrosis in the presence of transforming growth factor-ß1. PLoS One. 2020. 21; 15(4): e0231905. doi: 10.1371/journal.pone.0231905.

24. Stöhr R, Schuh A, Heine GH, Brandenburg V. FGF23 in Cardiovascular Disease: Innocent Bystander or Active Mediator? Front. Endocrinol 2018; 9: 351. Doi: 10.3389/fendo.2018.00351.

25. Vergaro G, Aimo A, Taurino E, Del Franco A, Fabiani I, Prontera C, Masotti S, Musetti V, Emdin M, Passino C. Discharge FGF-23 level predicts one year outcome in patients admitted with acute heart failure. Int J Cardiol. 2021. 1; 336: 98–104. Doi: 10.1016/j.ijcard.2021.05.028.

26. Eggers KM, Lindhagen L, Baron T, Erlinge D, Hjort M, Jernberg T, Marko-Varga G, Rezeli M, Spaak J, Lindahl B. Predicting outcome in acute myocardial infarction: an analysis investigating 175 circulating biomarkers. Eur Heart J Cardiovasc Care. 2021. 1;10(7):806-812. Doi: 10.1093/ehjacc/zuaa014.

27. Liu M, Xia P, Tan Z, Song T, Mei K, Wang J, Ma J, Jiang Y, Zhang J, Zhao Y, Yu P, Liu X. Fibroblast growth factor-23 and the risk of cardiovascular diseases and mortality in the general population: A systematic review and dose-response meta-analysis. Front Cardiovasc Med. 2022. 3; 9: 989574. doi: 10.3389/fcvm.2022.989574

28. Heleen Binnenmars S, Hoogslag GE, Yeung SMH, Brouwers FP, Bakker SJL, Van Gilst WH, Gansevoort RT, Navis G, Voors AA, De Borst MH. Fibrobast Growth Factor 23 and Risk of New Onset Heart Failure With Preserved or Reduced Ejection Fraction: The PREVEND Study. J Am Heart Assoc. 2022. 2; 11(15): e024952. doi: 10.1161/JAHA.121.024952.

29. Gutiérrez-Landaluce C, Aceña Á, Pello A, Martínez-Milla J, González-Lorenzo Ó, Tarín N, Cristóbal C, Blanco-Colio LM, Martín-Ventura JL, Huelmos A, López-Castillo M, Alonso J, López Bescós L, Alonso-Pulpón L, González-Parra E, Egido J, Mahíllo-Fernández I, Lorenzo Ó, González-Casaus ML, Tuñón J. Parathormone levels add prognostic ability to N-terminal pro-brain natriuretic peptide in stable coronary patients. ESC Heart Fail. 2021 Aug;8(4):2713–2722. doi: 10.1002/ehf2.13331.

